# Professional grief among psycho-oncologists in Germany: A cross-sectional survey study

**DOI:** 10.1101/2025.08.11.25332944

**Authors:** Svenja Wandke, Klaus Lang, Martin Härter, Karin Oechsle, Carsten Bokemeyer, Mareike Rutenkröger, Isabelle Scholl

**Author notes:** Correspondence to: Svenja Wandke University Medical Center Hamburg-Eppendorf Department of Medical Psychology Martinistraße 52 Building W26 20246 Hamburg GERMANY. first author (Svenja Wandke). shared senior authorship (Mareike Rutenkröger, Isabelle Scholl).

## Abstract

**Background:** Professional grief refers to the emotional response healthcare professionals may experience following patient deaths. Although likely relevant in clinical practice, this phenomenon has been largely overlooked in research— particularly among psycho-oncologists. This study examined the emotional impact, coping strategies, and support needs related to professional grief in a German sample of psycho-oncologists.

**Methods:** A cross-sectional online survey was disseminated via professional associations and randomly selected cancer centers in Germany. Eligible participants were currently working in psycho-oncology and had experienced at least one patient death. The survey included two established instruments (Texas Revised Inventory of Grief – Present Feelings [TRIG-D], Professional Bereavement Scale [PBS-D]) and self-developed items on emotional responses, coping behaviors, and support needs.

**Results:** 258 participants (79% female; mean age 48 years) were included. Participants reported an average of three patient deaths per month and moderate overall distress (scale 0–10, M = 5.02, SD = 2.14). Scores on both grief scales were in the low to moderate range. Most participants even reported positive effects of professional grief on their sense of purpose and views on life and death, with minimal impact on relationships. Common coping strategies included adopting an accepting stance and peer support.

**Conclusion:** This study offers the first quantitative insight into professional grief among psycho-oncologists in Germany. Professional grief levels were low to moderate. The reported positive changes suggest that patient deaths may even pose a chance for professional growth. Future research should explore relevant risk and protective factors to guide targeted support.

## Background

Grief is commonly understood as a response to the loss of someone with whom a meaningful bond existed^1^. Traditional perspectives and research on grief predominantly focus on personal relationships rather than the professional ones between healthcare professionals (HCPs) and their patients^2^. In cancer care, patient loss is a frequent and presumably emotionally charged experience for HCPs^3^. Due to the nature of cancer care trajectories, HCPs may develop significant connections with patients over time^4^. Recent evidence suggests that grief experienced in the professional context—termed professional grief—differs from personal grief in important ways, though the boundary between the two is not always clear^2^. In a recent scoping review, we outlined core features of professional grief in oncology, including its frequent occurence, lower intensity, and confinement to the professional setting⁶. It may involve emotions such as guilt or a sense of responsibility and is often perceived as inappropriate within professional or societal norms^5^.

The impact of patient deaths on HCPs’ mental health and well-being can be considerable. Previous research links professional grief to adverse outcomes including declining mental health^6^, reduced overall well-being and quality of life^2,4,6,7^, as well as compromised patient care^8,9^. The majority of previous research has focused on nurses and physicians^5^. However, psycho-oncologists are also closely involved in multidisciplinary cancer care, in line with national and international guidelines^10–12^. Psycho-oncologists, by definition, provide psychosocial support to individuals facing serious illness and end-of-life situations^13^. Their work is grounded in sustained therapeutic relationships, often involving regular encounters over extended periods^14^. These emotionally intense interactions—frequently centered around existential concerns and psychological well-being—can foster strong interpersonal bonds that may increase vulnerability to professional grief^2,15^. Despite clinical importance and emotional complexity of their work, there is a marked lack of empirical data on professional grief among psycho-oncologists, particularly within European healthcare contexts. The existing literature is heavily skewed toward Anglo-American settings, and studies originating in Germany are almost absent^5^.

Few studies have used quantitative methods to examine professional grief, limiting the generalisability of findings^5^. To date, only one quantitative study has investigated professional grief among psycho-oncologists, based on an Israeli sample^2^, reporting moderate grief levels and a link between high grief (especially with low social acknowledgment) and compassion fatigue. In Germany, only one qualitative interview study^16^ has explored this topic, revealing both positive and negative effects of patient deaths, the importance of peer support, and unmet needs regarding emotional support and training. However, no study has yet addressed all three evidence gaps: (1) non-Anglo-American samples, (2) psycho-oncologists as a distinct group, and (3) quantitative data on professional grief. This cross-sectional study is the first to do so. It aims to explore the experiences, coping mechanisms, and unmet support needs of German psycho-oncologists facing patient loss.

## Methods

### Study Design

This study employed a cross-sectional design using an anonymous, one-time online survey. The study was conducted and reported in accordance with the STROBE (Strengthening the Reporting of Observational Studies in Epidemiology) guidelines^17^ for cross-sectional studies (supplementary file 1). Due to its’ heightened specificity to online survey studies, the CHERRIES (Checklist for Reporting Results of Internet E-Surveys) checklist^18^ was used additionally (supplementary file 2).

### Sampling and Recruitment

Participants were recruited via several relevant professional organizations (for a comprehensive list see supplementary file 3). These organizations distributed the survey invitation—containing a link to participate—via email and/or newsletter between August and September 2024. The survey remained open for participation from August until November 2024. To supplement this recruitment strategy, contact details of psycho-oncological services from cancer centers were identified using the online directory of cancer care facilities certified by the German Cancer Society (“Oncomap”). From this database, 180 facilities were randomly selected and invited to participate. These facilities were also encouraged to forward the invitation to colleagues within their networks. The random selection focused on centers treating cancer entities with most absolute numbers of deaths in Germany – i.e. lung cancer, pancreatic cancer and breast cancer^19^. For each cancer entity, every fifth center listed in “Oncomap” was chosen within each federal state in Germany.

As an incentive, participants were given the opportunity to enter a raffle for one of 20 online vouchers, each valued at €15.

The estimated total population of psycho-oncologists in Germany was derived from Schulz et al.^20^ and set at N = 3,183. Based on this figure, the minimum required sample size was calculated to be 93, assuming a margin of error (e) of 0.1, a proportion (p) of 0.5, and a z-score of 1.96. The formula used for this calculation was: 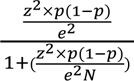. Inclusion criteria specified that participants had to provide psychosocial care to cancer patients and have experienced the death of at least one patient in the context of their professional role. Formal training as a psycho-oncologist by completing a course accredited by the German Cancer Association was not required for participation.

### Data collection

An open online survey was administered via LimeSurvey^21^. Before participation, all respondents provided electronic informed consent after being informed of the study’s voluntary and anonymous nature. The questionnaire was developed based on the authors’ prior research, including a scoping review on professional grief in cancer care^5^ and a qualitative interview study^16^. Relevant conceptual frameworks and existing instruments were integrated. In addition to standardized instruments, the survey included several self-developed items. The complete questionnaire is available in Supplementary File 4.

Section A of the survey focused on the experience of professional grief. In A1, participants were asked to rate the general level of distress they experienced in response to patient deaths, ranging from 0 = “not distressed at all” to 10 = “very distressed.” A2 assessed participants’ agreement with a series of statements concerning their emotional and professional attitudes toward patient death. Items were rated on a 5-point Likert scale from 1 = “strongly disagree” to 5 = “strongly agree”.

To measure grief-related distress, we used the German adaptation of the Texas Revised Inventory of Grief – Present Feelings Subscale^22^, comprising 16 items (A3, Supplementary File 4). Responses were recorded on a 5-point Likert scale ranging from 1 = “completely false” to 5 = “completely true", with higher scores indicating greater emotional burden. The original instrument has demonstrated sound psychometric properties^22^, although no validated cut-off scores exist.

To further assess professional grief, we included the Professional Bereavement Scale^23^, which evaluates grief responses in HCPs. It comprises two subscales: Short-Term Bereavement Reactions (SBR; items 1–17) and Accumulated Global Changes (AGC; items 18–32), both in Section A4 of Supplementary File 4. Participants rated SBR items by recalling their most recent patient loss and assessing emotional reactions within the first week post-loss on a 5-point Likert scale from 0 = “not at all” to 4 = “extremely strong". For the AGC subscale, they indicated the extent of perceived long-term personal change using a 5-point scale from 0 = “No (no such change or the change was not induced by experiencing patient deaths)” to 4 = “Yes, a great deal". Since no validated German version of the PBS existed, the scale was translated and culturally adapted using the TRAPD methodology^24^. Psychometric validation is reported elsewhere.

In A5 participants reflected on their experienced most stressful patient death and rated the intensity of emotional responses (0 = “not at all” to 4 = “very intense”). A6 consisted of checklist-style items on contextual distress factors. A7, adapted from Delafontaine et al.^6^, assessed perceived impact of recent patient deaths across life domains (-2 = “strong negative impact” to 2 = “strong positive impact”). In A8, participants indicated how long such effects typically lasted (“a few moments to an hour” to “more than a year”).

Section B of the survey addressed how psycho-oncologists cope with patient deaths. In B1, participants rated the perceived importance of various coping strategies on a 5-point Likert scale from 1 = “not important at all” to 5 = “very important.” B2 focused on the personal relevance of different support figures in the coping process, using the same rating scale.

Section C focused on unmet needs related to coping with patient deaths. In C1, participants indicated their level of agreement with a series of statements about such unmet needs, using a 5-point Likert scale. C2 provided an opportunity for participants to elaborate on additional needs or provide further comments in open-ended text fields.

Section D gathered demographic information.

### Statistical analysis

All data were analyzed using IBM SPSS Statistics Version 29^25^. Item-level analyses included all participants who fulfilled the inclusion criteria. Descriptive statistics, including frequencies and percentages, were calculated at the item level. For scale-based analyses, only cases in which participants completed at least 70% of the items within a given scale were included, in accordance with best practice recommendations and the scales original authors^23,26,27^. A detailed overview of excluded and imputed cases per scale is provided in Supplementary File 5. For each included scale, descriptive statistics such as mean and standard deviation were computed.

To explore potential risk or protective factors, we performed descriptive post hoc subgroup analyses of overall distress and TRIG-measured grief. Subgroups were defined based on (1) the number of colleagues participants work with and (2) their years of professional experience. These variables were selected due to the known relevance of peer support^5,16,28^ and prior research focusing on novice HCP, stating professional inexperience as a potential risk factor^28–30^. Given the exploratory nature and uneven subgroup sizes, findings are interpreted descriptively.

## Results

### Sample characteristics

A total of N = 289 individuals accessed the survey link. Of these, 30 participants were excluded due to reasons such as lack of informed consent or failure to meet inclusion criteria (for a detailed overview, see figure 1, flow of participants). Additionally, we excluded one case due to implausible and incomplete response patterns. Thus, N = 258 participants were deemed eligible for analysis.

**Figure 1.**
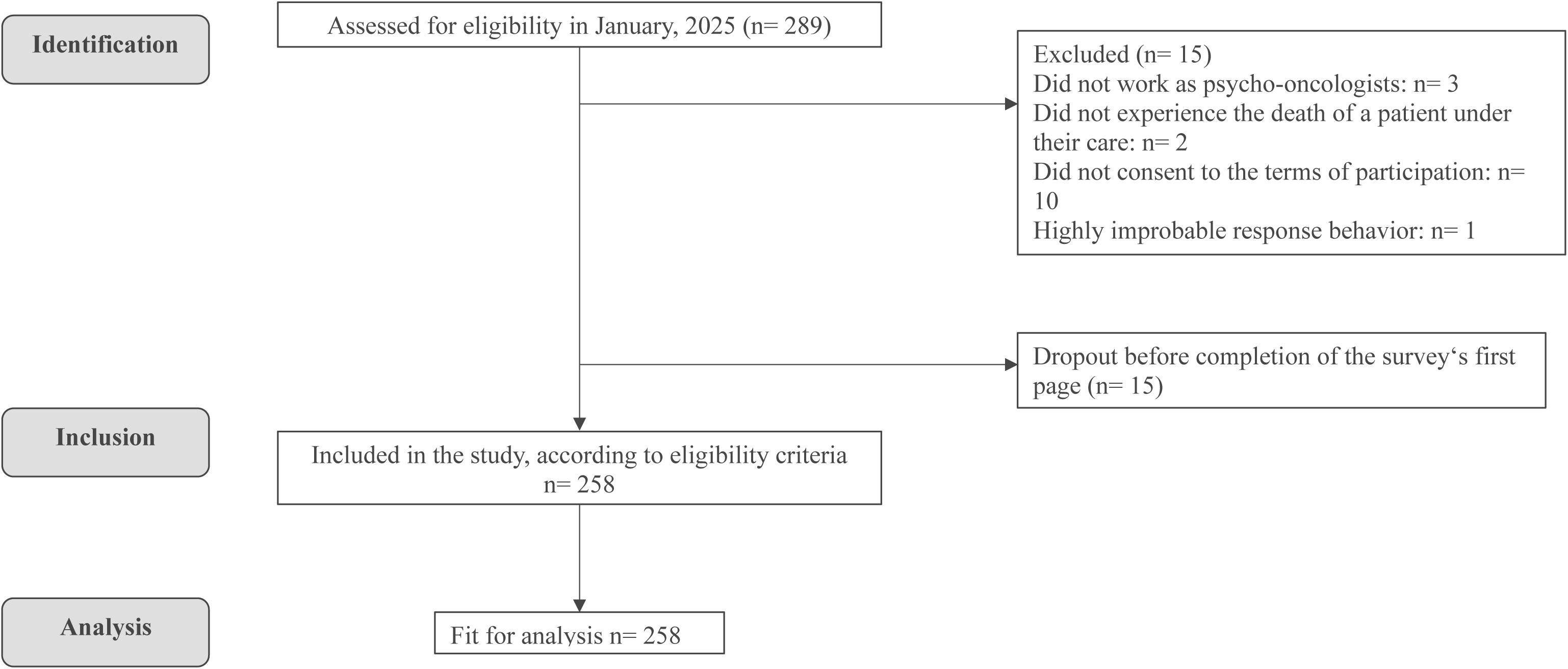
Flow of participants.

A detailed overview of sample characteristics is presented in Table 1. Participants had a mean age 48 years, were predominantly female and reported caring for M ≈ 36 patients per month (range: 0–200), while experiencing M ≈ 3 patient deaths per month (range: 0–25).

**Table 1.**
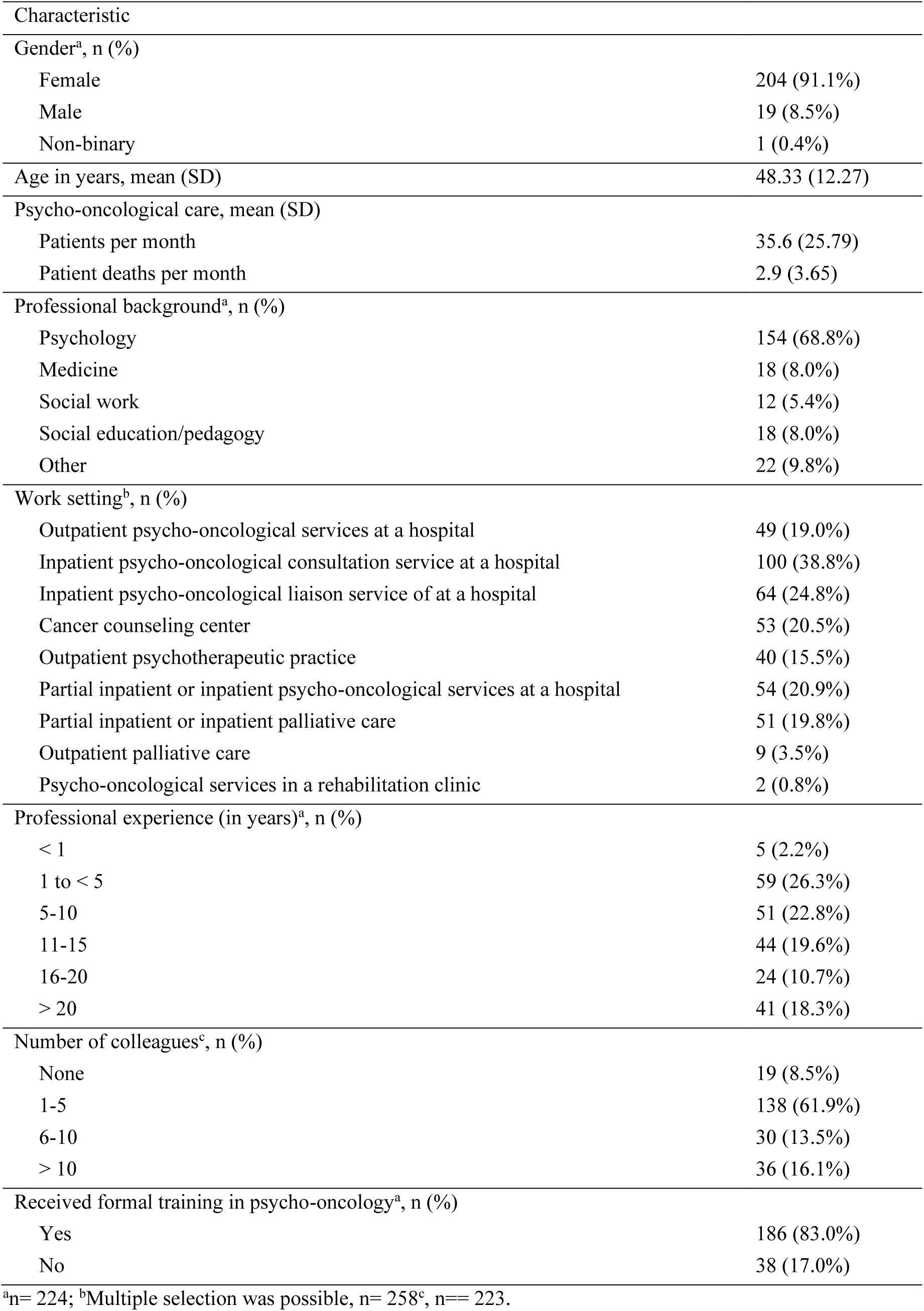
Sample characteristics.

### The experiences of professional grief

#### Distress and emotional response following patient deaths

Overall, participants reported a moderate level of distress related to patient deaths, with an average rating of M = 5.02 (SD = 2.14) on a scale from 1 ("not distressed at all") to 10 ("very distressed"). This finding is further supported by the grief-related measures. Although no validated cut-off scores are currently available for the PBS or TRIG, the observed values can be cautiously interpreted in relation to the possible score ranges, suggesting overall low to moderate levels of professional grief in this sample. With a mean sum score of M = 18.54 (SD = 8.52; range [17–85]), short-term bereavement reactions measured by the appropriate subscale of the PBS appeared rather limited. Similarly, low levels of grief were observed using the TRIG – Present Feelings Subscale, with a mean score of M = 24.65 (SD = 6.17; range [16–80]). However, the second subscale of the PBS revealed moderate levels of accumulated global changes, with a mean score of M = 28.71 (SD = 11.01; range [0–60]).

The intensity of emotional responses following patient deaths varied considerably among participants (see Table 2). The emotions reported with the highest mean intensity were sorrow/regret (M= 2.74), grief (M= 2.27), sadness (M= 1.77), and helplessness (M= 1.57). Among these, helplessness showed the greatest variability, with responses distributed almost evenly across all levels of intensity, indicating a wide range of individual experiences (SD = 1.23). In contrast, emotions such as indifference (M= .15), shame (.21) or guilt (.24) were reported as either not experienced or experienced at low intensity by most participants (> 90%).

**Table 2.** Emotional impact. Ranking of the emotional impact of patient deaths, with emotions rated as most intensely felt being at the top

| Emotion | Mean<br>(SD) | Not at all<br>(0) | A Little<br>(1) | Some<br>(2) | Intense<br>(3) | Very<br>Intense<br>(4) |
| --- | --- | --- | --- | --- | --- | --- |
| Frequencies (%) |  |  |  |  |  |  |
| Sorrow/ Regret | 2.74<br>(0.92) | 5<br>(2.1%) | 16<br>(6.7%) | 62<br>(25.9%) | 110<br>(46.0%) | 46<br>(19.2%) |
| Grief | 2.27<br>(1.01) | 15<br>(6.3%) | 31<br>(13.0%) | 89<br>(37.2%) | 83<br>(34.7%) | 21<br>(8.8%) |
| Sadness | 1.77<br>(1.05) | 29<br>(12.1%) | 64<br>(26.8%) | 90<br>(37.7%) | 44<br>(18.4%) | 12<br>(5.0%) |
| Helplessness | 1.57<br>(1.23) | 64<br>(26.8%) | 51<br>(21.3%) | 60<br>(25.1%) | 52<br>(21.8%) | 12<br>(5.0%) |
| Relief | 1.36<br>(1.01) | 60<br>(25.1%) | 65<br>(27.2%) | 84<br>(35.1%) | 28<br>(11.7%) | 2<br>(.8%) |
| Overwhelm | 1.18<br>(1.15) | 85<br>(35.6%) | 71<br>(29.7%) | 45<br>(18.8%) | 30<br>(12.6%) | 8 (3.3%) |
| Apprehen-<br>sion/Concern | 1.00<br>(1.02) | 101<br>(42.3%) | 65<br>(27.2%) | 46<br>(19.2%) | 25<br>(10.5%) | 2<br>(.8%) |
| Satisfaction | .88<br>(1.00) | 115<br>(48.1%) | 55<br>(23.0%) | 52<br>(21.8%) | 16<br>(6.7%) | 1<br>(.4%) |
| Fright | .83<br>(1.09) | 123<br>(51.5%) | 66<br>(27.6%) | 26<br>(10.9%) | 15<br>(6.3%) | 9 (3.8%) |
| Anger | .69<br>(1.00) | 139<br>(58.2%) | 59<br>(24.7%) | 21<br>(8.8%) | 16<br>(6.7%) | 4 (1.7%) |
| Despair | .39 (.75) | 175<br>(73.5%) | 40<br>(16.8%) | 17<br>(7.1%) | 5 (2.1%) | 1<br>(.4%) |
| Guilt | .24 (.61) | 199<br>(83.3%) | 25<br>(10.5%) | 13<br>(5.4%) | 1<br>(.4%) | 1<br>(.4%) |
| Shame | .21 (.58) | 205<br>(86.1%) | 17<br>(7.1%) | 14<br>(5.9%) | 2<br>(.8%) | 0<br>(0%) |
| Indifference | .15 (.48) | 212<br>(88.7%) | 19<br>(7.9%) | 6 (2.5%) | 2<br>(.8%) | 0<br>(0%) |

n= 239

#### Factors influencing the degree of perceived distress

Drawing on prior research, this study explored specific patient characteristics, psycho-oncologist traits, and aspects of the therapeutic relationship that may intensify distress following the death of a patient. Several factors were identified by the majority of participants (>50%) as contributing to heightened emotional burden. These included situations in which the deceased patient had *underage children* (71.3%), *a long duration of the therapeutic relationship* (67.8%), *young age of the deceased patient* (66.7%), or the *relationship was experienced as especially intense or deep* (64.7%). Additional distress was reported when participants identified themselves strongly with the patient due to *personal similarities*, such as similar age or background (58.5%), or when the *patient’s death was perceived as particularly agonizing* (57.8%). In contrast, the timing or manner in which psycho-oncologists were informed about a patient’s death was not seen as a contributor to distress by most participants (see Supplementary File 6, table 1).

#### Impact across different domains and duration of effects related to patient deaths

Most participants reported that their exposure to patient deaths had little to no effect on their leisure activities (68.9%), patient relationships (62.3%), and personal relationships (57.5%) (see Supplementary File 6, table 2). In contrast, the majority indicated that patient deaths had a positive or a strong positive impact on their sense of purpose (74.1%), their personal representation of life (77.2%) or death (63.5%). Nevertheless, a notable subgroup of 64 participants (28%) reported negative consequences in at least one of all assessed domains. Regarding the duration of these effects, the majority of participants (80.2%) indicated that the impact of patient deaths lasted no longer than one week, with 32.2% indicating impacts lasting a few hours up to a day and 40.5% stating to perceive the impact of patient deaths a few days up to a week. Only a very small proportion (1.3%) reported that the consequences persisted for more than one year (see Supplementary File 6, table 3).

**Table 3.** Coping strategies. Ranking of strategies applied by participants to cope with patient deaths, with strategies rated as most important being at the top

| Coping Strategy | Mean (SD) | Not im-<br>portant at<br>all<br>(1) | Rather un-<br>important<br>(2) | Neither im-<br>portant nor<br>unimportant<br>(3) | Rather im-<br>portant<br>(4) | Very im-<br>portant<br>(5) |
| --- | --- | --- | --- | --- | --- | --- |
| Frequencies (%) |  |  |  |  |  |  |
| Adopting an accepting stance towards the death of people | 4.57 (.62) | 1 (.4%) | 1 (.4%) | 6 (2.7%) | 78 (34.7%) | 139 (61.8%) |
| Sharing with colleagues (informal, e.g. during the lunch break) | 4.53 (.76) | 2 (.9%) | 6 (2.7%) | 7 (3.1%) | 65 (28.9%) | 145 (64.4) |
| Commemorating the deceased patient | 4.34 (.68) | 1 (.4%) | 2 (.9%) | 14 (6.2%) | 110 (48.9%) | 98 (43.6%) |
| Sharing with colleagues (formal, e.g. as part of super- or intervision) | 4.24 (.90) | 1 (.4%) | 15 (6.7%) | 19 (8.4%) | 85 (37.8%) | 105 (46.7%) |
| Expression of own emotions (emotional relief) | 4.23 (.77) | 0 (0%) | 8 (3.6%) | 22 (9.8%) | 106 (47.1%) | 89 (39.6%) |
| Finding a positive balance (e.g. vacations, spending time with friends, reading happy books) | 4.17 (1.04) | 8 (3.6%) | 13 (5.8%) | 18 (8.0%) | 80 (35.6%) | 106 (47.1%) |
| Reflecting on the topics of death/dying | 4.07 (.91) | 3 (1.3%) | 13 (5.8%) | 29 (12.9%) | 100 (44.4%) | 80 (35.6%) |
| Exercise (e.g. sport, yoga or going for a walk) | 3.98 (1.09) | 7 (3.1%) | 23 (10.2%) | 23 (10.2%) | 86 (38.2%) | 86 (38.2%) |
| Dealing with one's own mortality | 3.96 (1.01) | 4 (1.8%) | 20 (8.9%) | 34 (15.1%) | 89 (39.6%) | 78 (34.7%) |
| Taking a break from work | 3.95 (1.00) | 5 (2.2%) | 17 (7.6%) | 38 (16.9%) | 90 (40.0%) | 75 (33.3%) |
| Farewell rituals (e.g. lighting a candle) | 3.72 (1.24) | 17 (7.6%) | 24 (10.7%) | 38 (16.9%) | 72 (32.0%) | 74 (32.9%) |
| Physical distance from the workplace | 3.56 (1.32) | 27 (12.0%) | 26 (11.6%) | 27 (12.0%) | 84 (37.3%) | 61 (27.1%) |
| Exchange in a private environment | 3.26 (1.06) | 11 (4.9%) | 52 (23.1%) | 49 (21.8%) | 94 (41.8%) | 19 (8.4%) |
| Distraction (e.g. reading or watching TV) | 3.26 (1.21) | 25 (11.1%) | 29 (12.9%) | 71 (31.6%) | 63 (28.0%) | 37 (16.4%) |
| Individual supervision | 3.22 (1.22) | 22 (9.8%) | 43 (19.1%) | 60 (26.7%) | 63 (28.0%) | 37 (16.4%) |
| Faith | 2.79 (1.43) | 63 (28.0%) | 38 (16.9%) | 38 (16.9%) | 56 (24.9%) | 30 (13.3%) |

n= 239

### Coping with professional grief

Overall, participants rated the proposed coping mechanisms as important (see Table 3). On a scale from 1 (not important at all) to 5 (very important), the highest-rated strategies were: *adopting an accepting attitude toward death* (M = 4.57), *informal exchange with colleagues* (M = 4.53), *commemorating the deceased patient* (M = 4.34), *formal exchange with colleagues* (M = 4.24), *expression of one’s own emotions* (emotional relief; M = 4.23), *finding a positive balance* (M = 4.17), *reflection on death and dying* (M = 4.07), and *exercise* (M = 3.98). *Faith* was the overall lowest rated coping strategy (M = 2.79), even though approximately a third of all participants reported that faith is an important or very important coping strategy when faced with patient deaths.

Regarding sources of support, 92% of participants considered colleagues to be at least an important resource in coping with patient deaths, with more than half (55.6%) even rating them as very important. Partners (68%) and friends (47.3%) were also commonly perceived as important sources of support. In contrast, 66.2% of respondents rated spiritual care providers as rather unimportant or not important at all in coping with patient deaths (see Supplementary File 6, table 4).

**Table 4.**
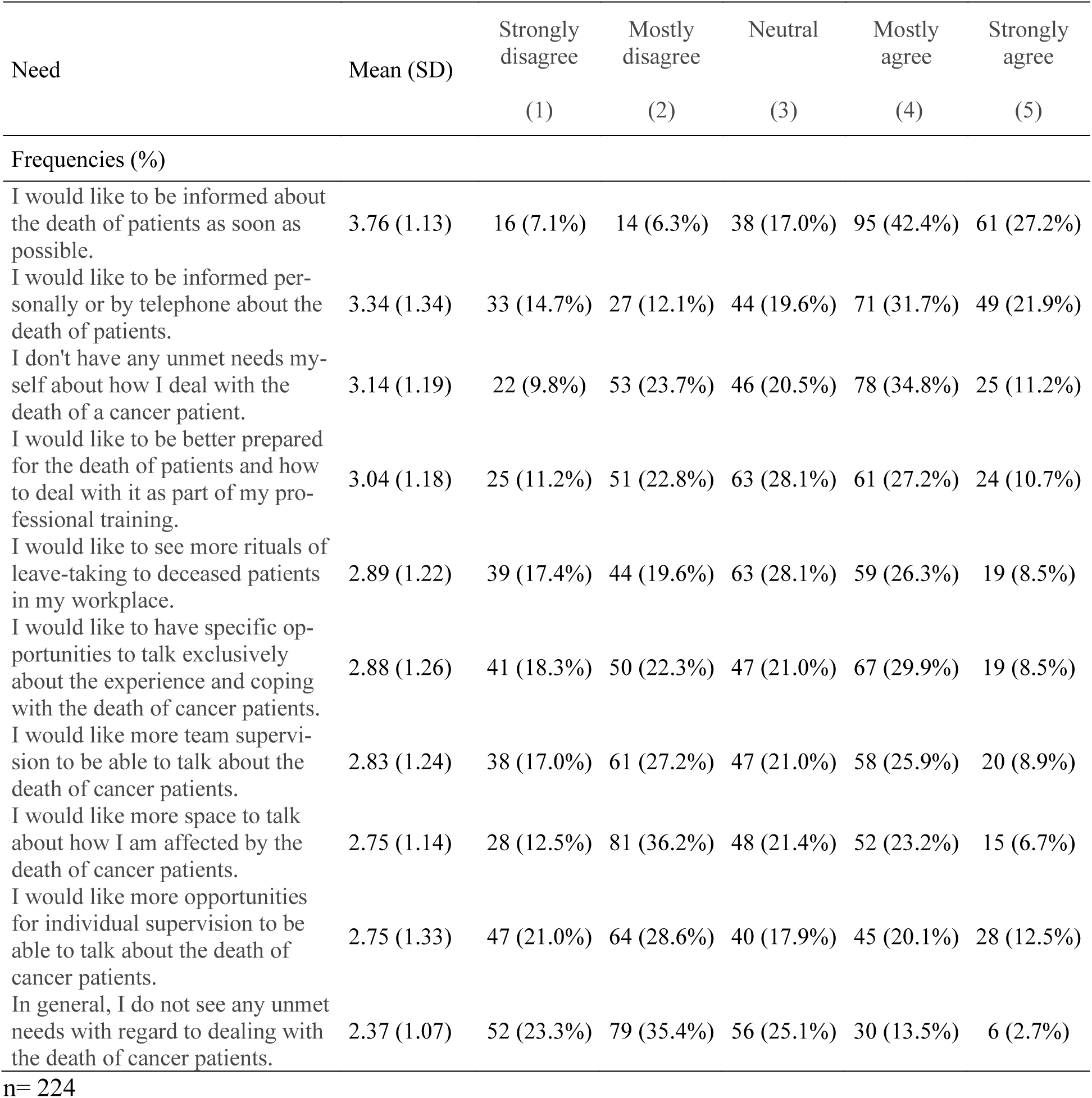
(Unmet) needs. Ranking of needs in regard to coping with patient deaths, with statements most agreed with being at the top

### Psycho-oncologists needs and preferences related to patient deaths

Participants’ responses showed high variability regarding their needs to effectively cope with patient deaths, with answers distributed across the full range of possible response options. However, two notable trends were identified: 69.6% of participants expressed a desire to be *informed about patient deaths as soon as possible*, and 53.6% indicated a preference for being *informed personally or via telephone*.

Overall, 58.7% of participants disagreed with the statement, “In general, I do not see any unmet needs with regard to dealing with the death of cancer patients,” suggesting that the majority perceived the opposite to be true and recognized at least some unmet needs. Furthermore, 64% did not only recognize unmet support needs in the psycho-oncological community but specifically reported having unmet support needs themselves. Regarding professional training, 37.9% of participants expressed a desire to be better equipped for handling patient deaths.

### Results of subgroup analysis

With respect to number of colleagues, only minor differences in grief and distress were observed (see Supplementary File 6, Table 5, Figures 1 and 2). A slight U-shaped trend emerged for grief, with psycho-oncologists reporting either no colleagues or more than ten colleagues showing higher TRIG scores. The highest distress levels were observed in those with more than ten colleagues.

In terms of professional experience (Supplementary File 6, Table 5, Figures 3 and 4), grief and distress levels varied but did not follow a linear pattern. The highest levels of grief and distress were reported by those with less than one year of experience. Interestingly, respondents with 11 to 15 years of experience reported the lowest grief levels, but the highest distress ratings.

## Discussion

The present findings suggest that psycho-oncologists experience professional grief as a natural and generally adaptive emotional response to patient deaths. The emotions reported—such as sorrow, sadness, grief, and occasionally helplessness—are emotionally similar to personal grief, though the low to moderate levels of grief assessed with the TRIG-D indicate that the intensity is lower than in personal grief^22^. Descriptively, the level of grief reported in our sample was somewhat lower than that observed in the Israeli sample investigated by Engler-Gross et al.^2^, potentially suggesting differences in experience or contextual factors. Previous findings on the impact of professional grief in healthcare professionals have been mixed ^4–6,28^, with some studies highlighting predominantly negative effects and others noting potential for growth. In our sample of psycho-oncologists, the experience of professional grief was not reported alongside long-term or global negative effects on personal or professional life. In fact, many participants reported positive psychological outcomes, particularly with respect to their views on life, death, and their sense of purpose.

Emotions such as guilt, which are commonly reported among physicians^5^, played a negligible role in the responses of psycho-oncologists. This divergence may be attributable to the distinct nature of their professional responsibilities. Unlike physicians, psycho-oncologists do not make medical decisions or bear the burden of treatment side effects or outcomes, which may shield them from feelings of moral responsibility or failure. These differences suggest that while professional grief is a common phenomenon across healthcare professions, its emotional expression and psychological consequences may vary depending on role-specific factors. Further comparative studies across disciplines are warranted to clarify such distinctions.

Coping mechanisms reported by participants were diverse, yet consistently rated as important. The presence of colleagues—both in informal and formal settings— emerged as a central support structure. Colleagues were not only identified as the most important source of support, but could also facilitate emotional relief, rated as the most important coping strategy. Compared to other healthcare professions, psychological disciplines may be generally more oriented toward emotional self-reflection and the open expression of one’s own emotional responses, for instance through established formats such as clinical supervision^31–34^. It is possible that this professional culture and training foster a greater readiness to acknowledge and process difficult emotions following patient deaths. This, in turn, might serve as a protective factor—potentially contributing to the comparatively lower levels of distress observed in this sample relative to findings from studies involving physicians or nurses^4,7^. While this interpretation is plausible, it requires further investigation to be substantiated. Nevertheless, these findings underscore the value of peer-based coping and highlight the potential for institutions to promote collegial exchange through regular supervision, peer discussion rounds, or the creation of informal spaces for supportive interaction.

In addition to social coping strategies, individual practices such as adopting an accepting attitude toward death and commemorating deceased patients were also perceived as highly beneficial. These coping mechanisms do not require the presence of others and may be particularly important in promoting self-regulated adaptation to loss.

The positive impacts reported by many participants may be understood in light of the concept of post-traumatic growth—defined as positive psychological change following the struggle with a traumatic event^35^. While it is most likely that none to few patient deaths may be experienced as traumatic, some encounters can be deeply moving or even destabilizing for individual professionals. The fact that many psycho-oncologists reported constructive outcomes from their experiences with death—such as strengthened values or deeper perspectives on life—suggests that professional grief may, in some cases, be a path to growth rather than distress. This interpretation aligns with research showing that perceived social support, feeling connected to others and reflective training can foster post-traumatic growth^36–39^. Viewing patient deaths as unavoidable distressing events, that yet yield an opportunity for post-distress growth may offer a useful framework: while grief responses are typically adaptive, insufficient coping resources or particularly distressing circumstances may still give rise to negative effects. As with post-traumatic stress and growth, these reactions may represent two sides of the same coin.

### Clinical implications

From a clinical standpoint, fostering collegial support structures should be a key priority within psycho-oncological and wider healthcare settings. Both formal (e.g., supervision) and informal (e.g., peer conversations) avenues should be actively promoted to support healthy grief processing. Given that individual practices such as reflection and rituals also play a role, healthcare institutions may benefit from integrating grief-sensitive approaches into their workplace culture—such as designated spaces for remembrance or optional farewell rituals. Additionally, the expressed need for better training in how to deal with patient deaths indicates that professional grief should be explicitly addressed within psycho-oncology, or more general: psychology and psychotherapy curricula, and continuing education.

### Research implications

On a research level, further studies are needed to explore potential risk factors for maladaptive or unprocessed professional grief. Identifying subgroups of professionals who may be more vulnerable—due to role-specific, personal, or contextual factors—could help inform targeted support measures. Moreover, understanding protective factors present among psycho-oncologists could guide interventions aimed at supporting other healthcare providers exposed to repeated patient loss. Our exploratory subgroup analyses suggest that grief and distress levels may vary with both professional experience and the number of colleagues, though no consistent pattern emerged. This warrants further and more thourough investigation. Developing validated tools and norm values for assessing professional grief would also be essential to advance this field methodologically.

### Strenghts and limitations

This study is the first to explore professional grief among psycho-oncologists in Germany using quantitative methods and one of the few to do so in a European context. Strengths include a national sample with varied professional experience and work settings, as well as the combined use of validated instruments and context-sensitive, self-developed items. However, several limitations must be noted.

First, the convenience sample may be subject to self-selection bias and may not represent all psycho-oncologists—particularly those less connected to professional networks or working in different institutional contexts. Although the recruitment strategy was broad, we did not contact the full population of approximately 3,200 psycho-oncologists in Germany. The reach of the invitation remains unclear, as mailing list sizes were unknown and forwarding was encouraged but not trackable. Self-selection may also occur earlier, as professionals who enter psycho-oncology might differ from others in their attitudes toward death and dying. Moreover, individuals experiencing severe professional grief—either within psycho-oncology or psychology more broadly—may have opted out due to stigma or fears of appearing “unprofessional,” despite guaranteed anonymity^2^.

Second, the absence of validated cut-off scores for the grief measures limits the interpretability of the reported low-to-moderate scores.

Third, while participants reported the average number of patient deaths per month, we did not assess how frequently individual patients were seen. As nearly 68% of respondents rated the length of the therapeutic relationship as relevant, this unmeasured variable could be a key factor influencing grief severity.

Finally, generalizability beyond the German context may be limited due to structural differences in psycho-oncological care. Future research should aim to replicate and expand on these findings internationally.

## Conclusion

This study offers a more in-depth empirical insight into the phenomenon of professional grief among psycho-oncologists in Germany. The findings suggest that, for most, professional grief is an emotionally manageable and even constructive experience. While distressing in the short term, patient deaths may even contribute to personal and professional growth. It remains unclear whether the experience of growth following patient deaths is primarily enabled by sufficient coping resources or influenced by other individual or contextual factors; further research is needed to clarify these pathways. At the same time, a substantial proportion of participants reported unmet needs, particularly related to support and training. These results highlight the importance of institutional structures that validate grief, facilitate peer exchange, and provide education on coping with loss. Further research is needed to better understand the variability in grief responses across healthcare roles and to develop evidence-based strategies for supporting healthcare professionals in emotionally demanding fields.

## Supporting information

Supplementary file 1, STROBE checklist

Supplementary file 2, CHERRIES checklist

Supplementary file 3, professional associations

Supplementary file 4, questionnaire

Supplementary file 5, imputation protocol

Supplementary file 6, additional tables pertaining to the results section

## Data Availability

Data available on request from the authors.

## Authorship statement

IS is the responsible principle investigator of the study. IS and SW were involved in planning and preparation of the study. SW developed the questionnaire, while all authors critically discussed and revised the questionnaire. SW recruited participants and collected and analysed the data. All authors interpreted the results. SW wrote the first draft of the manuscript. KL, MH, KO, CB, MR and IS critically revised the manuscript for important intellectual content. All authors gave final approval of the version to be published and agreed to be accountable for the work.

## Data availability statement

Data available on request from the authors.

## Ethics statement

This study was carried out according to the latest version of the Helsinki Declaration of the World Medical Association and respecting principles of good scientific practice. The local Ethics Committee of the University Medical Center Hamburg-Eppendorf gave approval prior to investigation (approval number: LPEK-0614). Study participation was voluntary and no foreseeable risks for participants resulted from the participation in this study. Participants were fully informed about the aims of the study, data collection and the use of collected data and written informed consent was obtained.

