## Supplementary file 2, CHERRIES checklist for "Professional grief among psycho-oncologists in Germany: A cross-sectional survey study"

*"Improving the Quality of Web Surveys: The Checklist for Reporting Results of Internet E-Surveys (CHERRIES)"*[1]

Table 1, CHERRIES checklist

| Item Category | Checklist Item | Explanation | |
|---|---|---|---|
| **Design** | Describe survey design | Describe target population, sample frame. Is the sample a convenience sample? (In "open" surveys this is most likely.) | See page 5-7. |
| **IRB (Institutio-nal Review Board) appro-val and infor-med consent process** | IRB approval | Mention whether the study has been approved by an IRB. | See Ethics Statement (page 21). |
| | Informed consent | Describe the informed consent process. Where were the participants told the length of time of the survey, which data were stored and where and for how long, who the investigator was, and the purpose of the study? | See supplementary file 3. In an additional file participants were informed about data security aspects. A detailed data protection concept was developed prior to the beginning of data collection. For this study, anonymized data from the survey is stored in electronic form at the Institute and Department of Medical Psychology at the University Medical Center Hamburg-Eppendorf's access-restricted servers for 10 years. Only research group member's have access to these files. Only fully anonymized datasets are used and analyzed, while separately stored email addresses for the raffle are saved on access-restricted servers and deleted after study completion. |
| | Data protection | If any personal information was collected or stored, describe what mechanisms were used to protect unauthorized access. | See informed consent |
| **Development and pre-testing** | Development and testing | State how the survey was developed, including whether the usability and technical functionality of the electronic questionnaire had been tested before fielding the questionnaire. | See page 7-9 and supplementary file 4. Before opening the survey to the public an internal pilot test was conducted, to ensure technical functionality and usability. |

| | | | |
|---|---|---|---|
| **Recruitment process and description of the sample having access to the questionnaire** | Open survey versus closed survey | An "open survey" is a survey open for each visitor of a site, while a closed survey is only open to a sample which the investigator knows (password-protected survey). | Open survey (see page 7). |
| | Contact mode | Indicate whether or not the initial contact with the potential participants was made on the Internet. (Investigators may also send out questionnaires by mail and allow for Web-based data entry.) | See page 6. |
| | Advertising the survey | How/where was the survey announced or advertised? Some examples are offline media (newspapers), or online (mailing lists – If yes, which ones?) or banner ads (Where were these banner ads posted and what did they look like?). It is important to know the wording of the announcement as it will heavily influence who chooses to participate. Ideally the survey announcement should be published as an appendix. | See page 6. Wordings of the survey announcement can be requested via E-Mail. |
| **Survey administration** | Web/E-Mail | State the type of e-survey (eg, one posted on a Web site, or one sent out through e-mail). If it is an e-mail survey, were the responses entered manually into a database, or was there an automatic method for capturing responses? | See page 7.<br>Data was automatically captured through LimeSurvey[2]. |

| | | | |
|---|---|---|---|
| | Context | Describe the Web site (for mailing list/newsgroup) in which the survey was posted. What is the Web site about, who is visiting it, what are visitors normally looking for? Discuss to what degree the content of the Web site could pre-select the sample or influence the results. For example, a survey about vaccination on an anti-immunization Web site will have different results from a Web survey conducted on a government Web site | Does not apply. The survey link was hosted with limesurvey[2]. LimeSurvey is an open-source online survey platform that enables users to create, distribute, and analyze surveys and questionnaires. The survey link (and website) was only visited by parties interested in the survey, since there is no other possible utilization. |
| | Mandatory/voluntary | Was it a mandatory survey to be filled in by every visitor who wanted to enter the Web site, or was it a voluntary survey? | See page 7. |
| | Incentives | Were any incentives offered (eg, monetary, prizes, or non-monetary incentives such as an offer to provide the survey results)? | See page 6. |
| | Time/Date | In what timeframe were the data collected? | See page 6. The survey remained open for participation from August until November 2024. |
| | Randomization of items or questionnaires | To prevent biases items can be randomized or alternated. | There was no randomization of items. |
| | Adaptive questioning | Use adaptive questioning (certain items, or only conditionally displayed based on responses to other items) to reduce number and complexity of the questions. | There were only two adaptive questions, pertaining to the inclusion and exclusion criteria. If participants answered “no” to those, they were not able to answer the survey and forwarded to an ending-page. |
| | Number of Items | What was the number of questionnaire items per page? The number of items is an important factor for the completion rate. | There was a total of 174 items distributed unevenly on 15 screens. The ProQOL was displayed as a whole, therefore the maximum number of items per page was 30. |
| | Number of screens (pages) | Over how many pages was the questionnaire distributed? The number of items is an important factor for the completion rate. | There were 15 screens in total, of which 11 included survey items, including sociodemographic information, and 4 included study information, checking of inclusion criteria and the option to comment on the survey; survey quest-<br>ions were not evenly distributed on the screens. |

| | | | |
|---|---|---|---|
| | Completeness check | It is technically possible to do consistency or completeness checks before the questionnaire is submitted. Was this done, and if "yes", how (usually JAVAScript)? An alternative is to check for completeness after the questionnaire has been submitted (and highlight mandatory items). If this has been done, it should be reported. All items should provide a non-response option such as "not applicable" or "rather not say", and selection of one response option should be enforced. | Every question, except the open-ended questions and the question asking for participants' sex, was set so a "soft forced choice", meaning that participants were automatically informed via a pop-up that mandatory questions were not answered yet, however that they have the option to progress in the survey without answering the question(s) first. |
| | Review step | State whether respondents were able to review and change their answers (eg, through a Back button or a Review step which displays a summary of the responses and asks the respondents if they are correct). | Participants were able to review and change their answers through a Back Button. |
| **Response rates** | Unique site visitor | If you provide view rates or participation rates, you need to define how you determined a unique visitor. There are different techniques available, based on IP addresses or cookies or both. | We did not collect IP addresses or cookies to ensure maximum anonymity. Overall, n = 289 individuals accessed the survey link (s. page 10). |
| | View rate (Ratio of unique survey visitors/unique site visitors | Requires counting unique visitors to the first page of the survey, divided by the number of unique site visitors (not page views!). It is not unusual to have view rates of less than 0.1 % if the survey is voluntary. | Does not apply, see unique site visitors. |
| | Participation rate (Ratio of visitors who agreed to participate)/unique first survey page visitors | Count the unique number of people who filled in the first survey page (or agreed to participate, for example by checking a checkbox), divided by visitors who visit the first page of the survey (or the informed consents page, if present). This can also be called "recruitment" rate. | Does not apply, see unique site visitors. |

| | | | |
|---|---|---|---|
| | Completion rate (Ratio of<br>users who finished the the survey/users who agreed to participate | The number of people submitting the last questionnaire page, divided by the number of people who agreed to participate (or submitted the first survey page). This is only relevant if there is a separate "informed consent" page or if the survey goes over several pages. This is a measure for attrition. Note that "completion" can involve leaving questionnaire items blank. This is not a measure for how completely questionnaires were filled in. (If you need a measure for this, use the word "completeness rate".) | 258/289= .89 |
| **Preventing multiple entries from the same individual** | Cookies used | Indicate whether cookies were used to assign a unique user identifier to each client computer. If so, mention the page on which the cookie was set and read, and how long the cookie was valid. Were duplicate entries avoided by preventing users access to the survey twice; or were duplicate database entries having the same user ID eliminated before analysis? In the latter case, which entries were kept for analysis (eg, the first entry or the most recent)? | No measures were taken in order to prevent multiple entries from the same individual, in order to ensure maximal anonymity of participants. |

| | | | |
|---|---|---|---|
| | IP check | Indicate whether the IP address of the client computer was used to identify potential duplicate entries from the same user. If so, mention the period of time for which no two entries from the same IP address were allowed (eg, 24 hours). Were duplicate entries avoided by preventing users with the same IP address access to the survey twice; or were duplicate database entries having the same IP address within a given period of time eliminated before analysis? If the latter, which entries were kept for analysis (eg, the first entry or the most recent)? | No measures were taken in order to prevent multiple entries from the same individual, in order to ensure maximal anonymity of participants. |
| | Log file analysis | Indicate whether other techniques to analyze the log file for identification of multiple entries were used. If so, please describe. | No measures were taken in order to prevent multiple entries from the same individual, in order to ensure maximal anonymity of participants. |
| | Registration | In “closed” (non-open) surveys, users need to login first and it is easier to prevent duplicate entries from the same user. Describe how this was done. For example, was the survey never displayed a second time once the user had filled it in, or was the username stored together with the survey results and later eliminated? If the latter, which entries were kept for analysis (eg, the first entry or the most recent)? | Does not apply. |
| **Analysis** | Handling of incomplete questionnaires | Were only completed questionnaires analyzed? Were questionnaires which terminated early (where, for example, users did not go through all questionnaire pages) also analyzed? | See page 9. |

| | | | |
|---|---|---|---|
| | Questionnaire submitted with an atypical timestamp | Some investigators may measure the time people needed to fill in a questionnaire and exclude questionnaires that were submitted too soon. Specify the timeframe that was used as a cut-off point, and describe how this point was determined. | We did not record the time participants needed to fill in a questionnaire. |
| | Statistical correction | Indicate whether any methods such as weighting of items or propensity scores have been used to adjust for the non-representative sample; if so, please describe the methods. | No measures in this form were taken. |

2. Limesurvey GmbH. / LimeSurvey: An Open Source survey tool /LimeSurvey GmbH, Hamburg, Germany. URL http://www.limesurvey.org.
