## Supplementary file 3, professional associations for "Professional grief among psycho-oncologists in Germany: A cross-sectional survey study"

**Supplementary file 3, Professional associations within the recruitment process**

List of professional associations, who forwarded the study invitation to their members:

- Psycho-Oncology Working Group of the German Cancer Aid's Network of Comprehensive Cancer Centers (*Arbeitsgruppe Psychoonkologie des Netzwerks Onkologische Spitzenzentren – CCC*)
- State Consortium for Cancer Counselling Services (*Bundesarbeitsgemeinschaft für Krebsberatungsstellen e.V.*)
- Association for Continued Education in Psycho-Social Oncology (*Weiterbildung Psychosoziale Onkologie e.V.*)
- Psychosocial Working Group within the Society for Pediatric Oncology and Hematology (*Psychosoziale Arbeitsgemeinschaft in der Pädiatrischen Onkologie und Hämatologie - AG in der Gesellschaft für Pädiatrische Onkologie und Hämatologie e.V.*)
- German Cancer Association's Consortium for Psycho-Oncology (*Arbeitsgemeinschaft Psychosoziale Onkologie der Deutschen Krebsgesellschaft*).
- Addition: members of a regional network of psycho-oncologists in the Hamburg metropolitan area (*Psychoonkologie Treffen – POT*)
