## Supplementary file 4, questionnaire for "Professional grief among psycho-oncologists in Germany: A cross-sectional survey study"

Supplementary file 4, Questionnaire[#]
[#]This is a translated version of the original questionnaire, only used for publication, the original version is in German and available upon request. The translation process involved two researchers with proficient language skills (native in German and fluent in English), one translating primarily and the other one checking the first draft. Disagreements were resolved through discussion.

Dear participant,

With this nationwide study, we aim to explore the emotions and attitudes you may have experienced or developed in your professional career following the death of patients. We are also interested in how you cope with the death of patients. If you work in psycho-oncology and have accompanied at least one cancer patient who has passed away, we would be very grateful for your participation. You do not need to have completed formal psycho-oncological training to take part.

By participating, you can make an important contribution to understanding the impact of cancer patient deaths on professionals working in psycho-oncology. For this purpose, we ask you to complete the following anonymous questionnaire. It includes questions about your experiences and some personal information (e.g., age, gender). Completing the questionnaire takes approximately 20 minutes.

You will also have the opportunity to enter a raffle to win one of 20 gift vouchers worth €15. The personal information required for this will be stored separately from your questionnaire responses and cannot be linked to your answers. Providing this information is voluntary.

Your participation is voluntary. Your data will only be used if you consent to take part in the study. There are no disadvantages to you whether you choose to participate or not. You can decide at any time before submitting the questionnaire to withdraw without providing a reason—simply close the window. This also applies if you have already given your consent.

If you have any questions about the study, please contact our research associate Svenja Wandke.

If you have read and understood the information about the study and data protection and wish to participate in the questionnaire, we kindly ask you to provide your consent on the next page. Please save this PDF so that you can refer to it at any time to review what you have consented to. If you do not wish to participate, no further action is required.

**Psycho-oncologists experiences and coping with patient deaths**
The following survey is about your personal experience and how you deal with the death of cancer patients. There are no 'right' or 'wrong' answers. Please answer according to your personal opinion.

Furthermore, as part of this study, we would like to test the suitability of a new questionnaire to assess the impact of patient death on cancer care providers. For this purpose, it is necessary to ask you some questions in a very similar form several times. Therefore, we ask that you complete the questionnaire in its entirety. You will be making an important contribution!

To begin, we want to find out about the impact that the death of a cancer patient may have on you. We would also like to examine the emotions and attitudes that you may have experienced or developed in your professional career following the death of a patient.

**I1**

1. Do you work as a psycho-oncologist? (You do not need to have undergone training in psycho-oncology to take part in this survey. You can also take part in the survey if you only care for/counsel a few patients with cancer.)

Yes/No

2. Have you ever accompanied a patient with cancer who died in the course of care?

Yes/No

**A1**

1. How distressed are you in general when cancer patients, you have cared for, die?
   0 = not distressed at all; 10 = very distressed.

| 0 | 1 | 2 | 3 | 4 | 5 | 6 | 7 | 8 | 9 | 10 |
|---|---|---|---|---|---|---|---|---|---|---|
| ☐ | ☐ | ☐ | ☐ | ☐ | ☐ | ☐ | ☐ | ☐ | ☐ | ☐ |

**A2**

2. Please indicate the extent to which you agree with each of the following statements.

| | Strongly disagree | Mostly disagree | Neutral | Mostly agree | Strongly agree |
|---|---|---|---|---|---|
| The death of a patient touches me. | ☐ | ☐ | ☐ | ☐ | ☐ |
| I have the right to be distressed by the patient deaths. | ☐ | ☐ | ☐ | ☐ | ☐ |
| It is unprofessional to grieve for deceased patients. | ☐ | ☐ | ☐ | ☐ | ☐ |
| I can cry in front of colleagues when I feel distressed by the patient deaths. | ☐ | ☐ | ☐ | ☐ | ☐ |
| Frequent patient deaths make me less able to empathize with patients. | ☐ | ☐ | ☐ | ☐ | ☐ |
| By experiencing patient deaths, I have developed a routine for dealing with death and dying. | ☐ | ☐ | ☐ | ☐ | ☐ |
| Through my experiences with patient deaths, I approach the topics of death and dying in my private environment with more composure. | ☐ | ☐ | ☐ | ☐ | ☐ |
| External circumstances prevent me from caring for dying patients in a way that I personally consider appropriate. | ☐ | ☐ | ☐ | ☐ | ☐ |
| I had to provide care for dying cancer patients in a way that did not correspond to my moral values and beliefs. | ☐ | ☐ | ☐ | ☐ | ☐ |

**A3 – Texas Revised Inventory of Grief – Present feelings subscale (Examples)**

3. Please answer the following questions by clicking on the numbers that best reflect your **current** feelings about the death of a patient.

| | Comple-tely false | False | Un-decided | True | Comple-tely true |
|---|---|---|---|---|---|
| I still cry when I think of the person who died. | ☐ | ☐ | ☐ | ☐ | ☐ |
| I still get upset when I think about the person who died. | ☐ | ☐ | ☐ | ☐ | ☐ |
| I cannot accept this person's death. | ☐ | ☐ | ☐ | ☐ | ☐ |
| […] | | | | | |
| I feel partly responsible for the death of the deceased person.[a] | ☐ | ☐ | ☐ | ☐ | ☐ |
| I have strong feelings of guilt when I think of the person who has died.[a] | ☐ | ☐ | ☐ | ☐ | ☐ |
| I am still very angry about the death of the person who died.[a] | ☐ | ☐ | ☐ | ☐ | ☐ |

*[a]Note to the reader: Items are not part of the original TRIG questionnaire, but of the german validated version.*

**A4 – Professional Bereavement Scale – Short-term bereavement reactions (Examples)**

4. Please recall your **last experience** with the death of a patient and rate, on a scale from not at all to extremly strong, the intensity of your reactions **within one week after the death** of this patient.

| | Not at all | Weak | Medium | Strong | Extremely strong | Not applicable* |
|---|---|---|---|---|---|---|
| I felt sad. | ☐ | ☐ | ☐ | ☐ | ☐ | ☐ |
| I blamed myself. | ☐ | ☐ | ☐ | ☐ | ☐ | ☐ |
| I doubted the value of my occupation. | ☐ | ☐ | ☐ | ☐ | ☐ | ☐ |
| […] | ☐ | ☐ | ☐ | ☐ | ☐ | ☐ |

** Please select if the statement does not apply to you (e.g. if the patient's family is unknown)*

**Professional Bereavement Scale – Accumulated global changes (Examples)**

5. Now that you have answered some questions about the possible short-term effects of patient deaths, the following is about the long-term changes caused by patient deaths. Compared with times before you encountered your first patient death, **you might have changed due to the patient deaths you have experienced so far in your professional life**. Please rate the extent to which you have been changed by patient deaths in each of following aspects.

| | No* | Yes, few | Yes, some | Yes, a lot | Yes, great deal |
|---|---|---|---|---|---|
| I am more aware that death is inevitable. | ☐ | ☐ | ☐ | ☐ | ☐ |
| I feel fatigued by my job. | ☐ | ☐ | ☐ | ☐ | ☐ |
| I am better at coping with patient deaths. | ☐ | ☐ | ☐ | ☐ | ☐ |
| […] | ☐ | ☐ | ☐ | ☐ | ☐ |

*****No such change or the change was not induced by experiencing patient deaths.

**A5**

6. Not all patient deaths have to be equally stressful. Please think back to the most stressful patient death you have experienced in your work as a psycho-oncologist. How intensely did you feel the following emotions after this patient died?

| | Not at all | A Little | Some | Intense | Very Intense |
|---|---|---|---|---|---|
| Indifference | ☐ | ☐ | ☐ | ☐ | ☐ |
| Satisfaction | ☐ | ☐ | ☐ | ☐ | ☐ |
| Relief | ☐ | ☐ | ☐ | ☐ | ☐ |
| Helplessness | ☐ | ☐ | ☐ | ☐ | ☐ |
| Overwhelm | ☐ | ☐ | ☐ | ☐ | ☐ |
| Sorrow/ Regret | ☐ | ☐ | ☐ | ☐ | ☐ |
| Grief | ☐ | ☐ | ☐ | ☐ | ☐ |
| Sadness | ☐ | ☐ | ☐ | ☐ | ☐ |
| Apprehension/Concern | ☐ | ☐ | ☐ | ☐ | ☐ |
| Fright | ☐ | ☐ | ☐ | ☐ | ☐ |
| Anger | ☐ | ☐ | ☐ | ☐ | ☐ |
| Shame | ☐ | ☐ | ☐ | ☐ | ☐ |
| Guilt | ☐ | ☐ | ☐ | ☐ | ☐ |
| Despair | ☐ | ☐ | ☐ | ☐ | ☐ |
| Others: | | | | | |

**A6**

7. Which of the following factors can contribute to you experiencing the death of patients as more distressing?

❑ The patient is similar to me (e.g. similar age, similar profession, etc.).
❑ The patient died at a time when I was already emotionally distressed for personal reasons (e.g. due to the death of someone close to me).

❑ The patient was young.
❑ The patient had (underage) children.

❑ I have accompanied the patient over a long period of time.
❑ I perceived the professional relationship as particularly intense.

❑ The patient had to suffer what I considered to be an agonizing death.
❑ The patient's death was unexpected or took me by surprise.

❑ I was informed about the patient's death in a way that was inappropriate to me (e.g. through the digital patient file).

- ☐ I was informed about the patient’s death in a situation that was inappropriate to me (e.g. shortly before the start of a meeting or appointment).
- ☐ I was informed about the patient's death at a time that was inappropriate to me (e.g. a long time after the death).
- ☐ Others:

**A7 – Adapted from Delafontaine et al. (2024)[1]**

8. How strongly do you rate the impact that the confrontation with the death of patients within the last month has had on you personally with regard to the following aspects?

| | Strong negative impact | Negative impact | No impact | Positive impact | Strong positive impact |
|---|---|---|---|---|---|
| Patient relationships | ☐ | ☐ | ☐ | ☐ | ☐ |
| Personal relationships | ☐ | ☐ | ☐ | ☐ | ☐ |
| Leisure time | ☐ | ☐ | ☐ | ☐ | ☐ |
| Sense of purpose/feeling of useful at work | ☐ | ☐ | ☐ | ☐ | ☐ |
| Personal representation of life | ☐ | ☐ | ☐ | ☐ | ☐ |
| Personal representation of death | ☐ | ☐ | ☐ | ☐ | ☐ |

[1]Delafontaine, A. C., Anders, R., Mathieu, B., Salathé, C. R., & Putois, B. (2024). Impact of confrontation to patient suffering and death on wellbeing and burnout in professionals: a cross-sectional study. BMC Palliative Care, 23(1), 74.

**A8**

9. How long do you usually feel the effects of a patient's death?

- ☐ A few moments to an hour
- ☐ A few hours to a day
- ☐ A few days to a week
- ☐ More than a week to a month
- ☐ Several months to one year
- ☐ More than a year

**B1**

The following section is about how you deal with the death of cancer patients.

10. How important are the following coping mechanisms for dealing with the death of cancer patients?

| | Not important at all | Rather unimportant | Neither important nor unimportant | Rather important | Very important |
|---|---|---|---|---|---|

| | | | | | |
|---|---|---|---|---|---|
| Expression of own emotions (emotional relief) | ☐ | ☐ | ☐ | ☐ | ☐ |
| Commemorating the deceased patient | ☐ | ☐ | ☐ | ☐ | ☐ |
| Taking a break from work | ☐ | ☐ | ☐ | ☐ | ☐ |
| Exercise (e.g. doing sports, yoga or going for a walk) | ☐ | ☐ | ☐ | ☐ | ☐ |
| Sharing with colleagues (informal, e.g. during the lunch break) | ☐ | ☐ | ☐ | ☐ | ☐ |
| Sharing with colleagues (formal, e.g. as part of supervision) | ☐ | ☐ | ☐ | ☐ | ☐ |
| Individual supervision | ☐ | ☐ | ☐ | ☐ | ☐ |
| Sharing in one's private environment | ☐ | ☐ | ☐ | ☐ | ☐ |
| Adopting an accepting stance towards death | ☐ | ☐ | ☐ | ☐ | ☐ |
| Physical distance from the workplace | ☐ | ☐ | ☐ | ☐ | ☐ |
| Distraction (e.g. reading or watching TV) | ☐ | ☐ | ☐ | ☐ | ☐ |
| Finding a positive balance (e.g. vacations, spending time with friends, reading happy books) | ☐ | ☐ | ☐ | ☐ | ☐ |
| Faith | ☐ | ☐ | ☐ | ☐ | ☐ |
| Reflection on death/dying | ☐ | ☐ | ☐ | ☐ | ☐ |
| Reflection on one's own mortality | ☐ | ☐ | ☐ | ☐ | ☐ |
| Farewell rituals (e.g. lighting a candle) | ☐ | ☐ | ☐ | ☐ | ☐ |

**B2**

11. How important are the following people for you personally when dealing with the death of patients?

| | Not important at all | Rather unimportant | Neither important nor unimportant | Rather important | Very important |
|---|---|---|---|---|---|
| Colleagues | ☐ | ☐ | ☐ | ☐ | ☐ |
| Friends | ☐ | ☐ | ☐ | ☐ | ☐ |
| Partner | ☐ | ☐ | ☐ | ☐ | ☐ |
| Spiritual care provider | ☐ | ☐ | ☐ | ☐ | ☐ |
| Others:* | | | | | |

*If yes, please enter these people here. If you wish, you can write how important they are to you personally when dealing with the death of patients.

**C1**

We would now like to discuss possible needs that you personally perceive when dealing with the death of a cancer patient.

12. Please indicate the extent to which you agree with each of the following statements.

| | Strongly disagree | Mostly disagree | Neutral | Mostly agree | Strongly agree |
|---|---|---|---|---|---|
| In general, I do not see any unmet needs with regard to dealing with the death of cancer patients. | ☐ | ☐ | ☐ | ☐ | ☐ |
| I don't have any unmet needs myself about how I deal with the death of a cancer patient. | ☐ | ☐ | ☐ | ☐ | ☐ |
| I would like more space to talk about how I am affected by the death of cancer patients. | ☐ | ☐ | ☐ | ☐ | ☐ |
| I would like more opportunities for individual supervision to be able to talk about the death of cancer patients. | ☐ | ☐ | ☐ | ☐ | ☐ |
| I would like more team supervision to be able to talk about the death of cancer patients. | ☐ | ☐ | ☐ | ☐ | ☐ |
| I would like to have specific opportunities to talk exclusively about the experience and coping with the death of cancer patients. | ☐ | ☐ | ☐ | ☐ | ☐ |
| I would like to be informed personally or by telephone about the death of patients. | ☐ | ☐ | ☐ | ☐ | ☐ |
| I would like to be informed about the death of patients as soon as possible. | ☐ | ☐ | ☐ | ☐ | ☐ |
| I would like to be better prepared for the death of patients and how to deal with it as part of my professional training. | ☐ | ☐ | ☐ | ☐ | ☐ |
| I would like to see more rituals of leave-taking to deceased patients in my workplace. | ☐ | ☐ | ☐ | ☐ | ☐ |

**C2**

13. Do you have (further) unmet needs with regard to how you cope with patient deaths?

14. Do you have any other comments you would like to share with us about your experiences or coping with patient deaths?

---

**D**

You have almost made it! Finally, we would like to ask you to answer a few questions about yourself.

1. Which gender do you consider yourself to be?
   - ❑ Male
   - ❑ Female
   - ❑ diverse/ non-binary
   - ❑ No answer

2. How old are you?

3. In which setting do you work?
   *You can check several options*
   - ❑ Outpatient psycho-oncological care in a clinic
   - ❑ Psycho-oncological consultation service of a clinic
   - ❑ Psycho-oncological liaison service of a clinic
   - ❑ Cancer counseling center
   - ❑ Psychotherapeutic practice
   - ❑ Partial inpatient or inpatient psycho-oncological care
   - ❑ Partial inpatient or inpatient palliative care
   - ❑ Outpatient palliative care
   - ❑ Psycho-oncological care in a rehabilitation clinic

4. What did you study?
   - ❑ Psychology
   - ❑ Medicine
   - ❑ Social work
   - ❑ Social pedagogy
   - ❑ Other:______________(please specify)

5. Have you completed further training as a psycho-oncologist certified by the German Cancer Association?
   Yes/No

6. How many psycho-oncological colleagues do you have?
   - ❑ None
   - ❑ 1 - 5 colleagues
   - ❑ 6 - 10 colleagues
   - ❑ More than 10 colleagues

7. How many years of professional experience do you have as a psycho-oncologist?
   - ❑ Less than 1 year
   - ❑ 1 year to less than 5 years
   - ❑ 5-10 years
   - ❑ 11-15 years
   - ❑ 16-20 years

❑ More than 20 years

8. On average, how many patients do you provide psycho-oncological support to each month?

9. How many of the patients you provide psycho-oncological support to die each month on average?
   It is sufficient to give an estimate amount.
