## Supplementary file 5, imputation protocol for "Professional grief among psycho-oncologists in Germany: A cross-sectional survey study"

**Supplementary file 5, Exclusion and imputation protocol**

***Texas Revised Inventory of Grief – Present feelings subscale (TRIG)***

Three participants (n= 3) had missing values on the TRIG scale, with all 16 items unanswered. Consequently, these individuals were excluded from the corresponding analyses, resulting in a final sample of n= 255 for this scale.

***Professional Bereavement Scale***

***Short-term bereavement reactions (PBS-SBR)***

For nine participants 17 items were missing on the respective scale, exceeding the 30% threshold for missing data. As a result, these cases were excluded from the scale-based analyses due to insufficient information to calculate a person-specific mean score. For one participant, one item was missing (respectively no more than 30% missing data). Therefore, a person-specific mean was imputed for the missing values, in line with the predefined criteria, resulting in a final sample of n= 249 for this scale.

***Accumulated global changes (AGC)***

For 13 participants the number of missing responses on the respective scale exceeded the threshold of 30% (i.e., more than 5 items missing). Therefore, these cases were excluded from the scale-based analyses.

For one participant one item was missing and for another participant three items were (for both: respectively no more than 30% missing data). Accordingly, person-specific mean scores were imputed for the missing items, following the established procedure, resulting in a final sample of n= 245 for this scale.
