## Supplementary file 6, additional tables pertaining to the results section for "Professional grief among psycho-oncologists in Germany: A cross-sectional survey study"

Table 1, Ranking of factors elevating the level of distress perceived by psycho-oncologists following patient-deaths, with factors heightening distress the most being at the top

| Factors elevating distress | Agreement n (%) |
|---|---|
| The patient had (underage) children. | 184 (71.3%) |
| I have accompanied the patient over a long period of time. | 175 (67.8%) |
| The patient was young. | 172 (66.7%) |
| I perceived the professional relationship as particularly intense. | 167 (64.7%) |
| The patient is similar to me (e.g. similar age, similar profession, etc.). | 151 (58.5%) |
| The patient had to suffer what I considered to be an agonizing death. | 149 (57.8%) |
| The patient died at a time when I was already emotionally distressed for personal reasons (e.g. due to the death of someone close to me). | 128 (49.6%) |
| The patient's death was unexpected or took me by surprise. | 100 (38.8% |
| I was informed about the patient's death in a way that was inappropriate to me (e.g. through the digital patient file). | 57 (22.1%) |
| I was informed about the patient's death in a situation that was inappropriate to me (e.g. shortly before the start of a meeting or appointment). | 48 (18.6%) |
| I was informed about the patient's death at a time that was inappropriate to me (e.g. a long time after the death). | 41 (15.9%) |

n=258

Table 2, Ranking of the aspects of life affected by patient deaths, with aspects rated as most impacted being at the top

| Aspect | Mean (SD) | Strong negative impact (-2) | Negative impact (-1) | No impact (0) | Positive impact (1) | Strong positive impact (2) |
|---|---|---|---|---|---|---|
| Frequencies (%) | | | | | | |
| Sense of purpose/ feeling of useful at work | .86 (.76) | 1 (.4%) | 10 (4.4%) | 48 (21.1%) | 130 (57.0%) | 39 (17.1%) |
| Personal representation of life | .82 (.70) | 0 (0%) | 14 (6.1%) | 38 (16.7%) | 151 (66.2%) | 25 (11.0%) |
| Personal representation of death | .67 (.81) | 0 (0%) | 26 (11.4%) | 57 (25%) | 123 (53.9%) | 22 (9.6%) |
| Patient relationships | .35 (.53) | 0 (0%) | 4 (1.8%) | 142 (62.3%) | 80 (35.1%) | 2 (0.9%) |
| Personal relationships | .32 (.61) | 1 (.4%) | 12 (5.3%) | 131 (57.5%) | 82 (36.0%) | 2 (.9%) |
| Leisure activity | .10 (.63) | 1 (.4%) | 26 (11.4%) | 157 (68.9%) | 38 (16.7%) | 6 (2.6%) |

n= 228

Table 3, Duration of patient deaths's impact on psycho-oncologists

| Duration | Frequencies (%) |
|---|---|
| A few moments to an hour | 17 (7.5%) |
| A few hours to a day | 73 (32.2%) |
| A few days to a week | 92 (40.5%) |
| More than a week to a month | 33 (14.5%) |
| Several months to one year | 9 (4.0%) |
| More than a year | 3 (1.3%) |

n=227

Table 4, Ranking of (groups of) people relied upon for support in coping with patient deaths, with aspects rated as most important being at the top

| Source of support | Mean (SD) | Not important at all (1) | Rather unimportant (2) | Neither important nor unimportant (3) | Rather important (4) | Very important (5) |
|---|---|---|---|---|---|---|
| Frequencies (%) | | | | | | |
| Colleagues | 4.43 (.79) | 2 (.9%) | 7 (3.1%) | 9 (4.0%) | 82 (36.4%) | 125 (55.6%) |
| Partner | 3.70 (1.15) | 13 (5.8%) | 28 (12.4%) | 31 (13.8%) | 95 (42.2%) | 58 (25.8%) |
| Friends | 3.22 (1.18) | 20 (8.9%) | 46 (20.5%) | 52 (23.2%) | 76 (22.9%) | 30 (13.4%) |
| Spiritual care provider | 2.00 (1.14) | 109 (48.4%) | 40 (17.8%) | 48 (21.3%) | 24 (10.7%) | 4 (1.8%) |

n= 225

Table 5, Subgroup analysis

| | | n | Mean (SD) |
|---|---|---|---|
| Number of colleagues[a] | | | |
| TRIG | None | 19 | 26.32 (4.93) |
| | 1-5 | 138 | 24.02 (5.96) |
| | 6-10 | 30 | 24.17 (5.68) |
| | >10 | 36 | 26.19 (6.66) |
| Distress | None | 19 | 5.05 (1.78) |
| | 1-5 | 138 | 4.75 (2.10) |
| | 6-10 | 30 | 5.10 (2.23) |
| | >10 | 36 | 6.14 (1.94) |
| Professional experience[b] | | | |
| TRIG | < 1 | 5 | 26.40 (6.03) |
| | 1 to < 5 | 59 | 24.58 (6.57) |
| | 5-10 | 51 | 25.14 (6.99) |
| | 11-15 | 44 | 24.00 (5.09) |
| | 16-20 | 24 | 24.46 (5.89) |
| | > 20 | 41 | 24.61 (5.04) |
| Distress | < 1 | 5 | 5.40 (1.95) |
| | 1 to < 5 | 59 | 5.07 (2.10) |

| | | |
|---|---|---|
| 5-10 | 51 | 4.96 (2.01) |
| 11-15 | 44 | 5.48 (1.86) |
| 16-20 | 24 | 5.08 (2.02) |
| > 20 | 41 | 4.66 (2.57) |

[a]n= 223; [b]n= 224

Figure 1, Perceived distress in accordance to number of colleagues

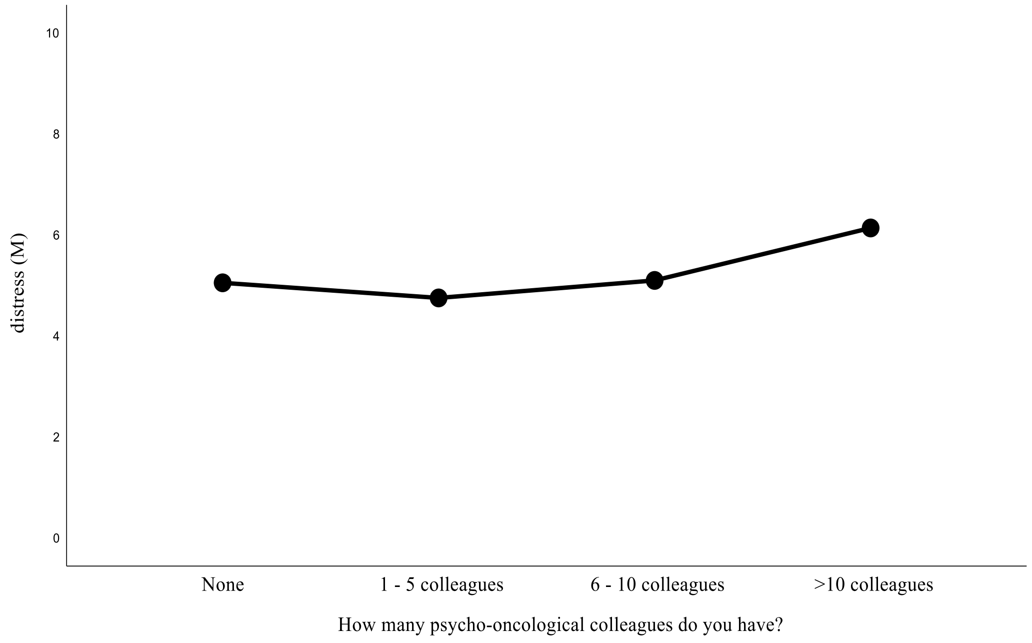


Figure 2, Grief in accordance to number of colleagues

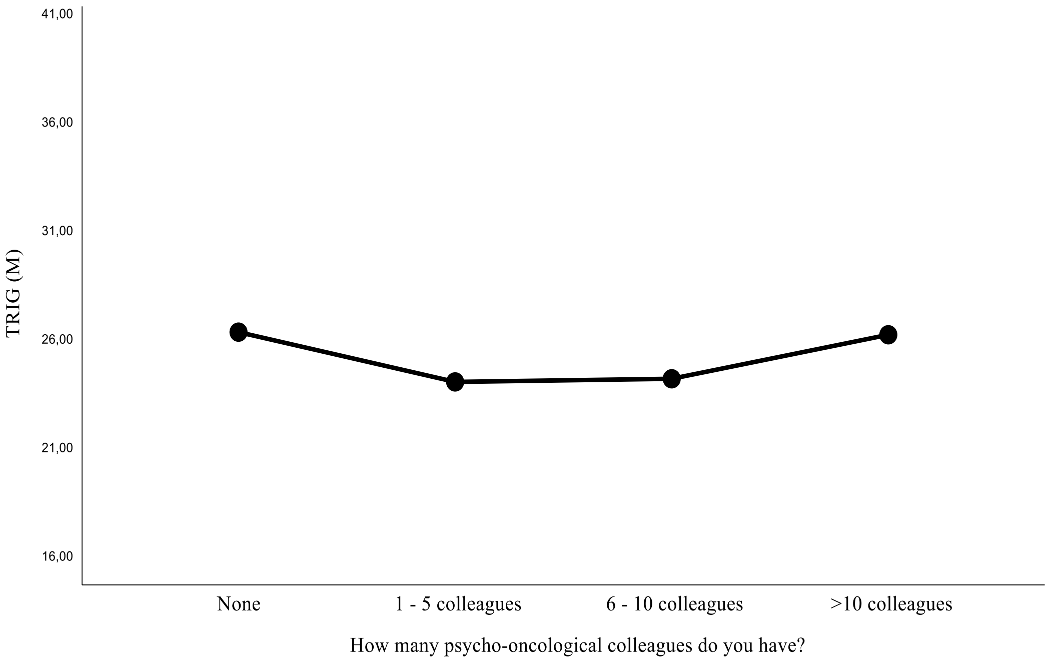

Figure 3, Perceived distress in accordance to years of professional experience

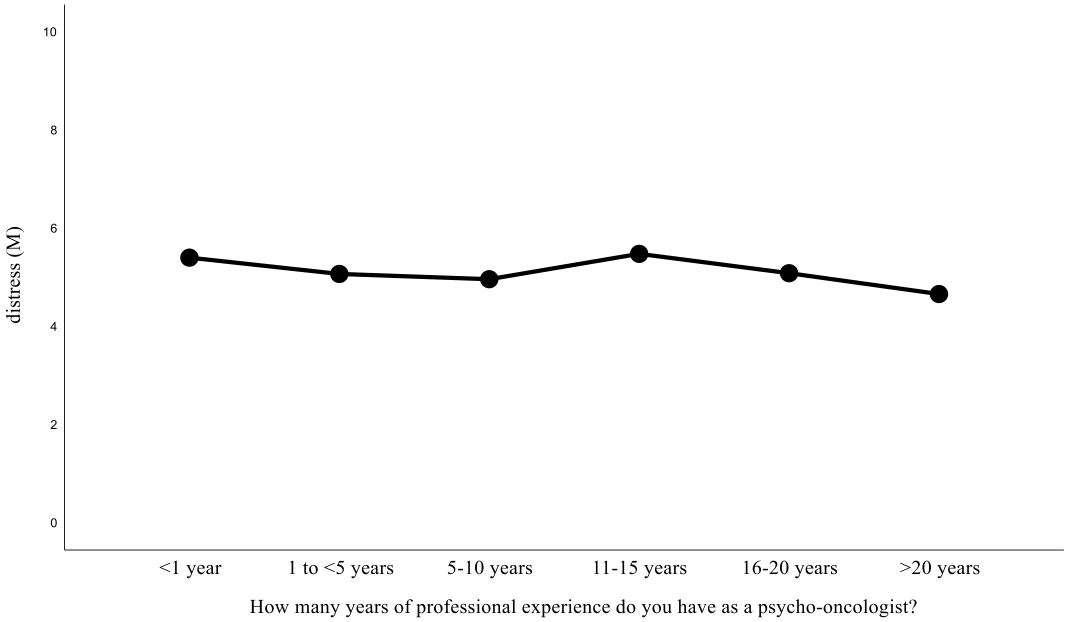


Figure 4, Grief in accordance to years of professional experience

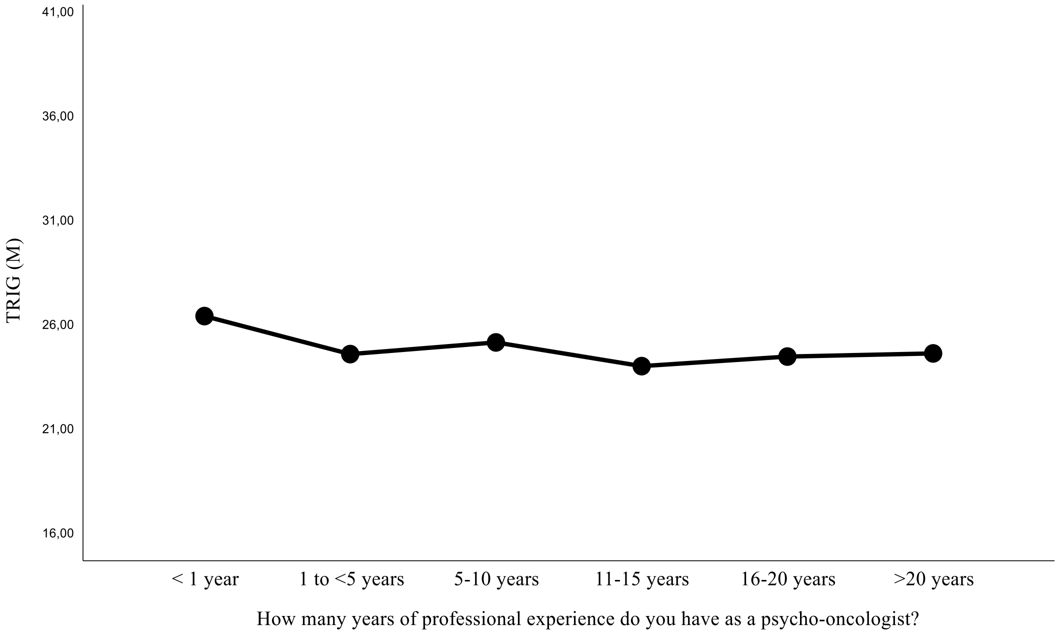
